# Neurology in a Changing Climate: A Scoping Review

**DOI:** 10.1101/2021.11.24.21266840

**Authors:** Shreya Louis, Alise Carlson, Abhilash Suresh, Joshua Rim, MaryAnn Mays, Daniel Ontaneda, Andrew Dhawan

## Abstract

**Importance:** Although the international community collectively seeks to reduce global temperature rise to less than 1.5ºC, there are already irreversible environmental changes that have occurred, and currently available evidence suggests these changes will continue to occur. As we begin to witness the effects of a warming planet on human health, it is imperative that as neurologists we anticipate the ways in which the epidemiology and incidence of neurologic disease may be affected.

**Objective:** In this review, we organize our analysis around three key themes related to climate change and neurologic health: extreme weather events and temperature fluctuations, emerging neuro-infectious diseases, and pollutant impacts. Across each of these key themes, we appraise and review recent literature relevant to neurological disease and the practice of neurology.

**Evidence Review:** Studies were identified using a set of relevant search terms relating to climate change and neurologic diseases in the PubMed repository for publications between 1990 and 2021. Studies were included if they pertained to human incidence or prevalence of disease, were in the English language, and were relevant to neurologic disease.

**Findings:** We identified a total of 136 articles, grouped into the three key themes of our study; extreme weather events and temperature fluctuations (23 studies), emerging neuro-infectious diseases (42 studies), and pollutant impacts (71 studies). Broadly, the studies included highlighted the relationships between neurologic symptom exacerbation and temperature variability, tick-borne infections and warming climates, and airborne pollutants and cerebrovascular disease incidence and severity.

**Conclusions and Relevance:** Our work highlights three key priorities for further work; namely, neuro-infectious disease risk mitigation, an understanding of the pathophysiology of airborne pollutants on the nervous system, and research into how to improve delivery of neurologic care in the face of climate-related disruptions.

**Key Points:** *Question:* How does climate change impact neurological disease?

*Findings:* 136 articles were identified relating neurologic disease to climate change. Articles were grouped into three key themes, highlighting evidence linking neurologic disease exacerbation to temperature fluctuation, tick-borne infections with climate change, and airborne pollutants to cerebrovascular disease.

*Meaning:* A substantial body of literature suggests that neurologic disease incidence, morbidity, and mortality is directly impacted by climate change and its sequelae, but much remains to be uncovered.

## Introduction

The effects of climate change on health are only beginning to be understood. In 2012, the Global Climate and Health Alliance drafted the Doha Declaration, a call to prioritize global policies to protect health as it is affected by climate change^1^. International calls for political advocacy and environmental justice surrounding the threat of climate change to human health have since followed.^2^ In September 2021, in the face of increasing extreme weather events attributed to climate change, over 220 medical journals published a joint editorial calling for the “urgent action to keep average global temperature increases below 1.5°C, halt the destruction of nature, and protect health”.^3^

Climate impacts on human health are well-documented^4–7^, but the impacts on patients with neurologic disease are less well characterized. The effects of a changing climate are unequally distributed and will disproportionately affect those in developing nations that have polluted less than wealthy nations.^8,9^ Climate change is also inextricably linked with airborne pollutant emissions produced by the combustion of fossil fuels, and studies on the topic have brought attention to the effects not only on the respiratory and cardiovascular systems^10–12^, but also neurological disorders^13–15^.

As the warming of our planet becomes increasingly apparent, there is an urgency to understand the impact of increasing temperatures on neurologic health in order to mitigate the effects on morbidity, mortality, and the burden on healthcare workers and health systems. Neurologists and neuroscientists have a duty not only to critically examine these potential changes but also to quantify their impact to better prepare patients and health care systems. Here, we present a scoping review of the literature pertaining to neurologic disease associated with climate change and airborne pollutants and identify avenues for further research in this increasingly important field.

## Methods / Literature Search strategy

Studies were identified by literature search of the PubMed repository for publications from January 1^st^ 1990 to May 30^th^ 2021 using the following search terms (alone or in combination with): “climate”, “climate change”, “global warming”, “air pollution/adverse effects”, “drought”, “flood”, “health systems”, “health policy”, “neurological disorders”, “neurology”, “multiple sclerosis”, “dementia”, “stroke”, “seizures”, “epilepsy”, headache”, or “migraine.” Search terms relating to specific disease entities were chosen due to their high global prevalence and burden of disease worldwide^16^.

Search results were input into the Covidence systematic review software where studies were screened, and duplicates were removed^17^. Authors SL and AD independently screened the remaining titles and study abstracts. Studies were excluded if the publications were without English translation, had primarily animal subjects (apart from neuroinfectious diseases in which case mosquito or tick-based studies were included), or if the study topic was not relevant to neurological conditions. The remaining studies were included for full-text review, categorization into the themes of the manuscript, and data extraction.

The Map depicting neuroinfectious diseases was created using the rnaturalearth v0.1^18^, rnaturalearthdata v0.1^19^, and ggplot2^20^ packages in R v4.1.1.^21^ Sankey diagram was created using the networkD3 v0.4^22^ package in R v4.1.1.^21^

## Results

Our search revealed a robust body of literature describing the effects of climate change in relation to neurologic disease (Figure 1). The included publications consisted of 136 studies with 42 studies from Asia, 41 studies from North America, 39 studies from Europe, 5 studies from Africa, 2 studies from Australia and Oceania, and 7 studies which spanned two or more continents. No relevant society guidelines, position statements, or governmental reports were identified.

**Figure 1:**
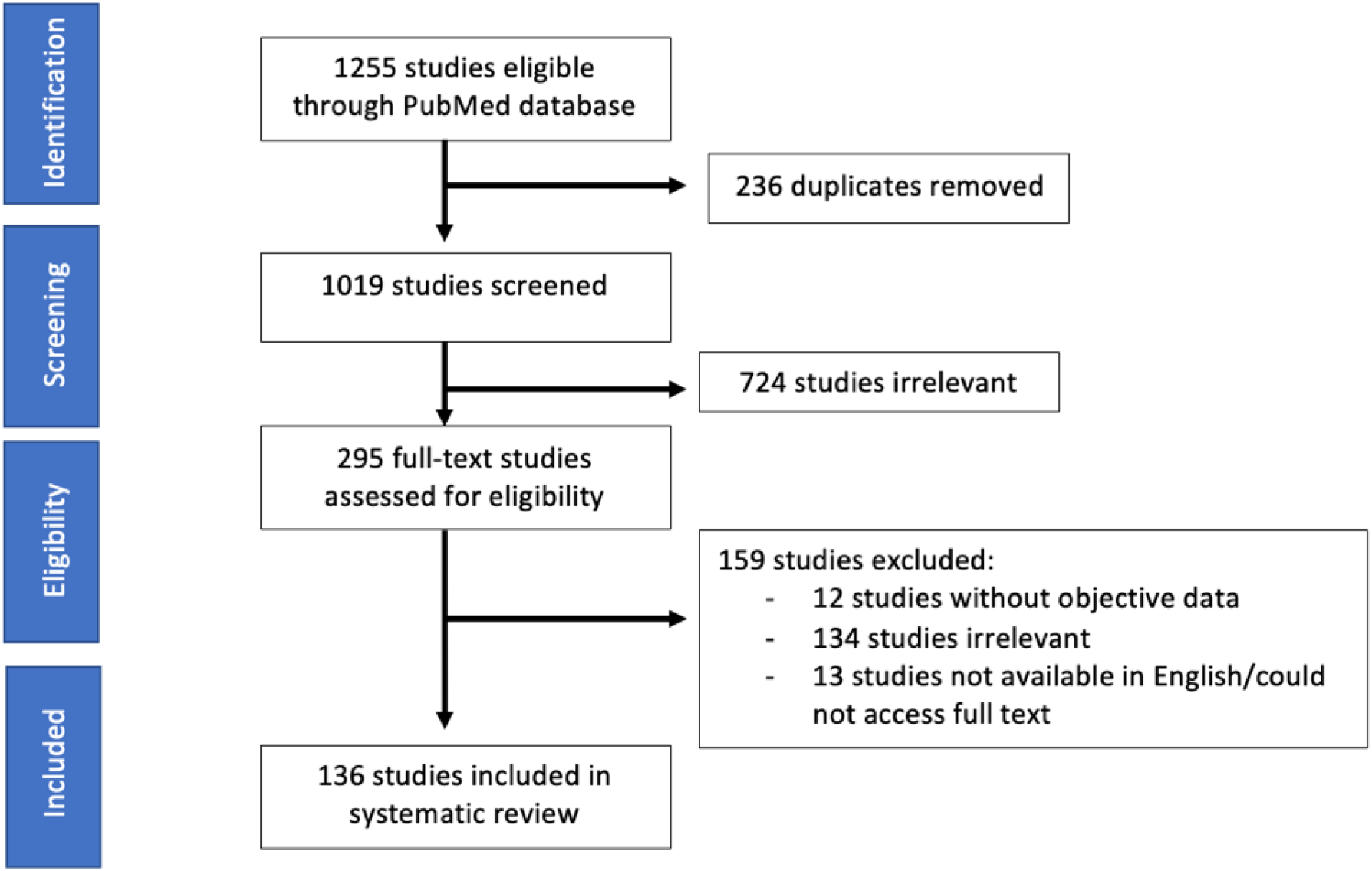
PRISMA diagram depicting studies included in this scoping review.

Two independent reviewers (SL and AD) grouped manuscripts into one of three categories: the effects of extreme weather and temperature changes on neurological conditions, emerging neuro-infectious diseases, and pollutant impacts. Relevant data including study population, intervention, comparators, and outcomes, were extracted from all studies and synthesized for this scoping review.

### The effects of temperature fluctuations on neurological conditions

#### Stroke

Our search identified 12 studies relating weather to ischemic stroke incidence, with the burden of evidence favoring an association of increased stroke in hotter weather as well as in extreme cold. We could not identify any consensus on mechanism relating changes in climate to ischemic stroke events. Six studies showed an increase in incidence of ischemic stroke events with increasing temperature and increased or decreased relative humidity.^23–28^ These results were in contrast to those from two large studies that identified increased stroke admissions with cold temperatures and hypothesized that colder temperatures contributed to stroke by inducing vasoconstriction and increasing blood viscosity.^29,30^ Bai and colleagues estimated the attributable fraction of stroke admissions in Ontario to cold temperature as 1.71%, with the burden of strokes attributable to moderate changes in temperatures as opposed to extreme changes.^29^ Temperature variability may also contribute to acute stroke incidence, as Lei and colleagues examined over 140,000 first-time stroke patients in Shenzhen, China, and attributed 2-4% of stroke cases to increased diurnal temperature range (greater than 5.5-8ºC in a 24h period, seasonally-dependent).^31^ The burden of disability-adjusted life years related to heat waves in a study by Yoon and colleagues in South Korea was found to be driven by cerebrovascular diseases, estimated at 72.1% of the total burden of disease.^32^ Two studies used climate projections into the mid-late 21^st^ century to predict ischemic stroke mortality, and showed an increase in years of life lost after accounting for population change, fertility, greenhouse gas emissions, and physical inactivity levels.^33,34^

#### Epilepsy

Our search strategy identified three studies relating the frequency of seizures in association with meteorological changes. Unstable weather, defined as alterations in barometric pressure by 10 hectopascals (hPa) and a change of 5ºC in 48h in a series of 30 patients with epilepsy, was associated with a season-dependent change in seizure frequency. Fluctuations were associated with changes in seizure frequency among 40% of participants in spring, autumn, and winter, but only 7% of participants in the summer.^35^ A study of 604 adult patients with epilepsy linked low atmospheric pressure and high relative air humidity to increased seizure risk but found that high ambient temperatures were associated with reduced seizure risk.^36^ Chiang and colleagues examined the number of outpatient and inpatient visits in patients with epilepsy as a function of air pollutants and ambient temperature and showed that there was a statistically significant association with ambient temperature and various air pollutants, with more visits during the winter months.^37^

#### Dementia

One study examined dementia-related hospital admissions and meteorological variables. Studies were retrospective observational cohorts, examined based on historical climate data. The largest such study analyzed data from 3,069,816 Medicare patients in New England over 10 years and used time-varying Cox proportional hazards models to estimate the association between hospital admissions for dementia and temperature variability. In the summer months, mean temperature increases of 1.5ºC associated with a 12% increased hazard of admission.^38^ Two studies showed that patients with dementia are often at higher risk of injury or death due to extreme heat events such as bushfires.^39,40^

#### Multiple Sclerosis

Three studies of patients with multiple sclerosis (MS) met the inclusion criteria for this study. While it is well-known that high temperatures exacerbate symptom burden in patients with MS, in one study, emergency department presentations for MS disease exacerbation (1,265 patients) were found to be associated with increased temperature variability on the preceding day. This study found an 8.81% increase in emergency department presentations per 1ºC increase in temperature range (95% CI 3.46-14.44%).^41^ An analysis of climatic variables and 260 MS admissions from Serbia showed that there was a statistically significant reduction of relapses in the period of high vitamin D exposure (a known geographical co-variate with disease incidence), and an increased number of relapses in the spring compared to other seasons.^42^ A randomized crossover study of MS patients undergoing physical therapy demonstrated differential responses in a warm climate (Spain) as opposed to a cold climate (Norway), with changes favoring warmer climates and persisting up to 6 months after treatment.^43^

#### Headache

Two studies examined headache in association with meteorological variables. A large study of over 22,000 headache visits to the emergency department showed that an increase in temperature by 5ºC was associated with a relative risk of headache presentation of 1.042 (95% CI 1.009 - 1.076).^44^ Mukamal and colleagues examined the associations between weather and headache incidence at a single center using a case-crossover design, examining temperature, barometric pressure, relative humidity, and pollutant levels in the 24-72h preceding presentation.^45^ In this study, higher temperatures were also associated with higher risk of presentation for headache, as did lower barometric pressure, particularly for non-migraine headache.^45^

### Emerging neuro-infectious diseases

We identified a robust body of literature related to climate patterns and neuro-infectious diseases, across 41 total studies, of which 10 were reviews and 31 were primary studies. Diseases studied included West Nile virus (WNV, 14 studies), Dengue virus (DENV, 2 studies), Japanese encephalitis (JEV, 8 studies), tick-borne encephalitis (TBE, 16 studies), and other emerging pathogens (Supplementary Table 1). Despite the growing literature on the subject, no studies were identified with our search strategy that related to COVID-19. Figure 2 highlights the geographic skew of evidence towards Europe and the significant paucity of studies in Africa and South America. The web of ecological interactions that leads to the emergence of these diseases and their transmission to humans is complex. Multiple factors, including climate, are implicated in disease transmission and prevalence. Several existing reviews highlighted the impact of extreme weather such as floods on the transmission of mosquito-borne and rodent-borne diseases. However, it is noted that a wide range of interactions including human population density and behavior, host-human interactions, land use patterns, pollutant effects, and disease adaptability all affect disease incidence.^46–49^

**Figure 2:**
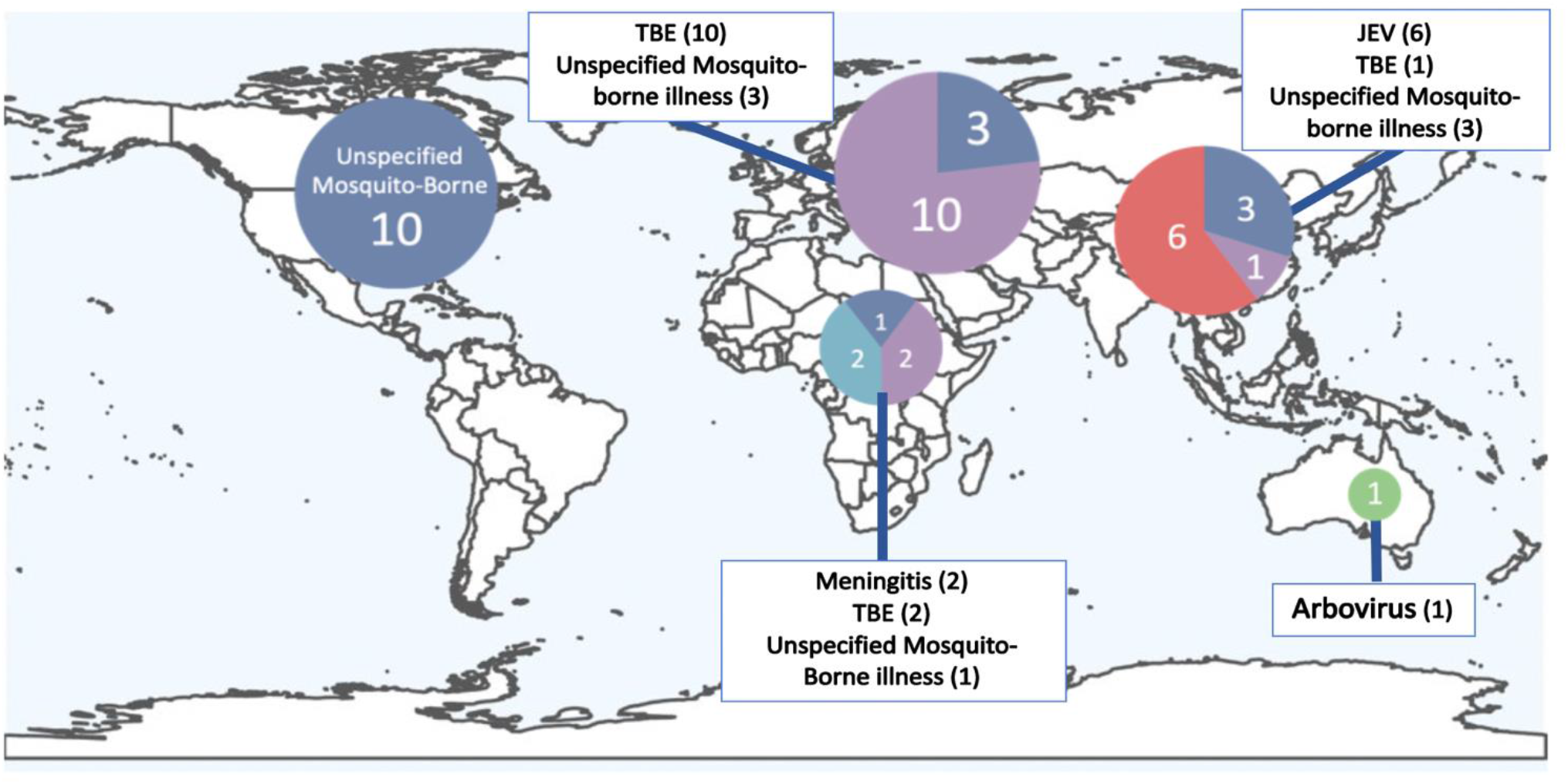
Neuro-infectious diseases studied by location. Map depicts number and type of reports for reports examining neuro-infectious diseases as stratified by location. 10 reports were published from North America, 13 reports from Europe, 5 reports from Africa, 10 reports from Asia, and 1 report from Australia. TBE refers to tick-borne encephalitis. JEV refers to Japanese encephalitis virus.

Most individual studies on the topic of climate associations with neuro-infectious diseases were observational, retrospective, and used vector incidence or disease prevalence with historical geospatial climactic data and time series modeling to establish associations between climate and infection risk. Infection risks are multifactorial and could often be explained by many factors; for instance, in examining why JEV incidence increased in the upslope regions of the Himalayas, Baylis and colleagues noted the incidence may be explained by either improved detection and inhomogeneous distribution of vaccines, an increased incidence of JEV due to climate change, or a combination.^50^ Our search also identified studies highlighting how regional factors beyond an increase in temperature could facilitate disease transmission. In the African ‘meningitis belt,’ aerosolized matter blown by the Harmattan winds is associated with increased meningitis transmission, whereas in the Czech Republic, flooding is associated with incidence of TBE.^51–53^ Similarly, reasons for the increase of TBE in Europe remain controversial, with some arguing that socioeconomic factors such as the fall of the Soviet Union caused changes in human behavior leading to increased exposure, and others arguing that climactic change better accounts for the increased incidence.^54,55^

We identified one mechanistic study where mathematical modeling of viral vector dynamics and climate variables predicted optimal disease transmission of WNV at temperatures of 23-26 degrees Celsius.^56^ Predictive modeling of WNV incidence in North America with climate simulations from 2021-2080 suggested an expansion of suitable conditions for the disease, particularly in the southern regions of the United States, due to higher temperatures, lower rainfall, a lengthening of mosquito season, and increase in droughts.^57–59^ Transmission periods for TBE in Europe are predicted to lengthen and new foci in Scandinavia may emerge, as suggested by analyses of epidemiological data between 1969-2018 from Sweden, East Germany, Czech Republic, Austria, Slovenia, and Northeast Italy.^60,61^

### Pollutant impacts

Studies relating pollutant impacts to neurologic disease were sub-grouped by disease: multiple sclerosis (MS), headaches, ischemic stroke and TIA, intracerebral hemorrhage (ICH), amyotrophic lateral sclerosis (ALS), Parkinson’s disease (PD), and dementia. Pollutants have also been shown to increase neurologic disease mortality in a general sense, with one study finding that periods of high air pollution due to heat waves and fires tripled the relative risk of death due to neurologic disease.^62^ Figure 3 summarizes the numerous interrelationships between pollutant exposures and neurologic disease.

**Figure 3:**
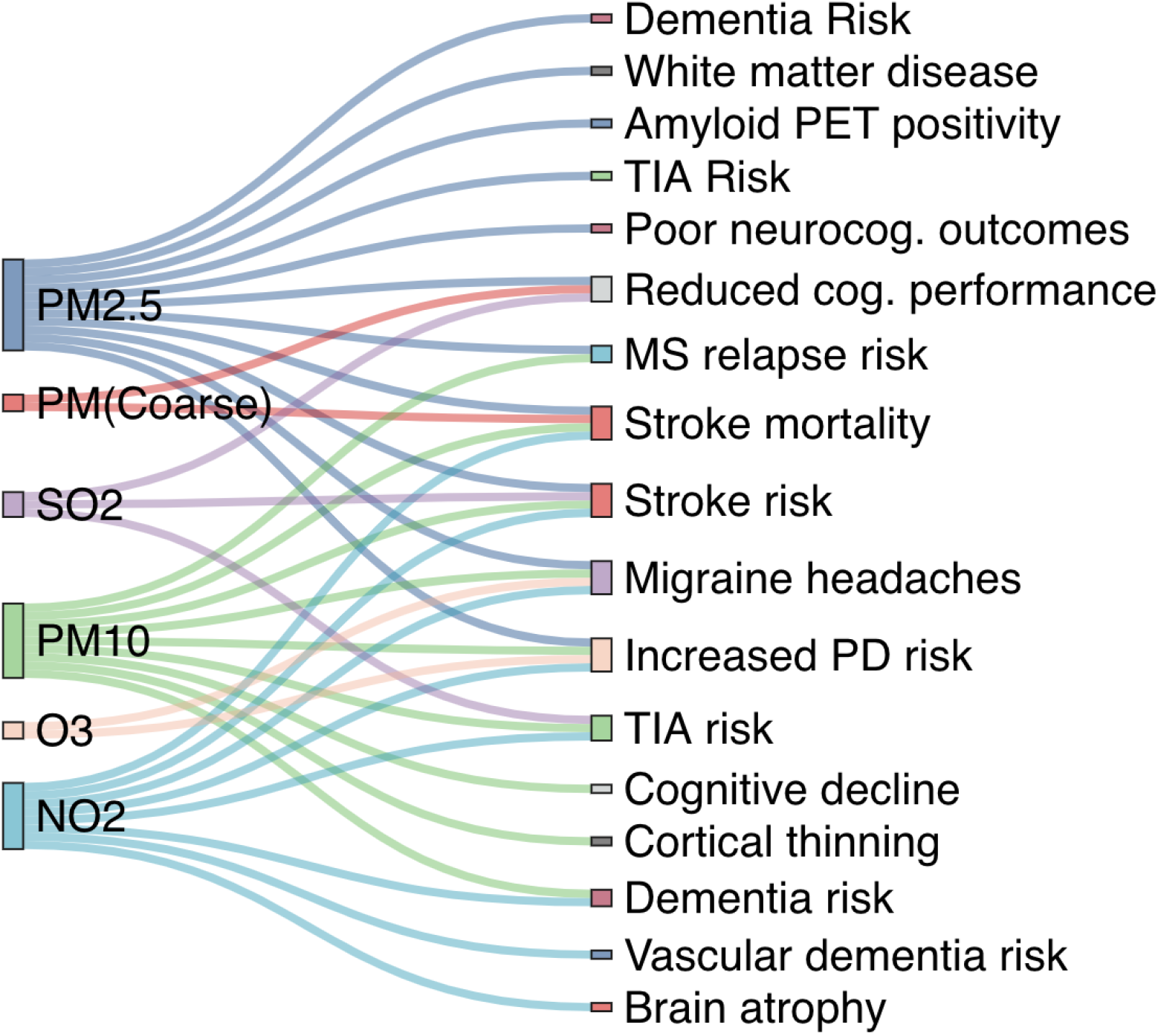
Relationships between pollutants and identified neurologic links. Sankey diagram depicting interrelationships between pollutant exposure (left) and neurologic outcome (right). PET refers to positron emissions tomography. TIA refers to transient ischemic attack. MS refers to multiple sclerosis. PD refers to Parkinson’s disease.

#### Dementia

The strongest environmental risk of air pollution was identified in the dementia literature, where nearly 6.1% of cases of incident dementia have been estimated to be attributable to PM2.5 (particulate matter with diameter less than 2.5 microns) and NO2 exposure.^63^ Clinical diagnosis of dementia^63,64^, and objective measures of cognitive decline, such as the MMSE^65,66^, semantic fluency, and word recall^67^ have been associated with prior years’ exposure to PM2.5. Indeed, it has been shown that amyloid PET scanning is more likely to be positive if living in an area with greater PM2.5 in the preceding 13-15 years.^68^ In patients who have pre-existing Alzheimer’s disease, Yitshak-shade and colleagues found association between PM2.5 exposure and risk of hospital admission.^69^ Imaging evidence of an association between PM2.5 exposure and neurodegenerative changes is provided by longitudinal studies of healthy patients with interval MRI scans, which have shown white matter loss in those exposed to greater concentrations of PM2.5.^70–74^ PM(coarse) has been less well-studied and only a single report linking exposure in women to decreased mini-mental status examination (MMSE) score has been published.^75^ Exposure to PM10, particularly in patients harboring the APOE4 allele, has been shown to associate both with dementia incidence and rate of cognitive decline, as well as front-temporal thinning on MRI.^66,74,76–78^ The effects of exposure to ground-level ozone (O3) remain less clear, as there have not been as strong associations with dementia incidence^76^, increased odds of amyloid PET positivity^68^, or MMSE score decline.^66^ Nitrogenous gases have shown an association with vascular dementia^79^, global brain atrophy^74^, and a slightly faster rate of decline in those with the APOE4 allele, with differing results for MMSE-scored cognitive decline and all-cause dementia.^63,64,66,75^ Sulfur dioxide (SO2) exposure has also been associated with MMSE score decline.^66,78^ Ten-year average exposure to black carbon has not shown an association with all-cause dementia.^64^

#### Ischemic stroke/Transient Ischemic Attack

Long-term exposures to airborne pollutants had substantial support for association with ischemic stroke. An analysis of the Global Burden of Disease study concluded that 9% of stroke disability-adjusted life years (DALYs) and 8.5% of stroke deaths could be attributed to PM2.5 exposure.^80^ After adjusting for sociodemographic factors, a study of 3,287 participants with incident stroke living within 100m of a roadway (associated with increased airborne pollutant exposure), was associated with a hazard ratio of 1.42 (95% CI 1.01-2.02) for ischemic stroke, though imaging-based biomarkers of small-vessel disease in a subset of these patients did not show significant association.^81,82^ A Chinese study of 12,291 ischemic strokes revealed a statistically significant association with PM1 and PM2.5 exposure three years prior to stroke event, but not with NO2 or PM10 exposure.^83^ Long-term exposures in a German study showed elevated risk of ischemic stroke regardless of particle size; examining PM2.5abs, PM2.5, and PM10.^84^ Long-term PM2.5 exposure examined in a study of 39,054 participants in China associated significantly with stroke mortality (HR 1.3, 95% CI 1.04-1.65).^85^

Studies examining short-term pollutant exposure and stroke risk generally used a case-crossover design. Results from these studies were conflicting and may depend on the population being studied, as well as the degree of geographic precision with which pollutant levels were examined. For example, of the four studies showing supportive evidence for short-term pollutant exposure to PM10 and stroke^86–89^, the largest studies Liu et al.^86^ and Yitshak-Sade^87^ had 279,890 events and 4,837 events respectively. On the other hand, two additional studies Royé et al.^30^ and Zheng et al.^90^ did not find significant associations between short-term PM10 exposure and stroke. Stroke mortality may also be worse on days with greater PM1, PM2.5, PM(coarse), and greater PM10.^91–93^ Short-term PM2.5 exposure was more controversial in its association with incident stroke and small vessel disease. We identified three clinical studies and one imaging-based study that did not reveal association^94–97^, and three clinical studies and one imaging-based study that did show association.^86,87,98,99^ Similar studies for first-time strokes and ozone exposure have shown mixed associations with a single study showing positive association^100^ and four studies showing no association.^30,96,100–102^ NO2 levels may also be associated with greater incidence of stroke-related admissions and stroke mortality in the short term,^89,103^ though two other studies did not show association.^30,90^ Shen and colleagues examined short-term SO2 exposure in relation to the rates of ischemic and hemorrhagic stroke and found a positive association,^104^ but two other studies did not find any association^30,90^ It may therefore be the case that individual factors, such as ethnic or genetic factors or degree of exposure, contribute significantly to the variability in this association.^95,98,101^

Only one study examining TIA risk and pollutants was identified. PM2.5, PM10, CO, NO2, and SO2 were significantly greater on days with TIA events than days without, and O3 did not show this association.^105^

#### Intracerebral hemorrhage

We identified three primary studies that examined risk of intracerebral hemorrhage attributable to environmental pollutants. In one study of 368 cases of small-vessel attributed ICH, short term exposure to increased PM2.5 was greater on the three days preceding the hemorrhage compared to a control period 15-17 days prior to the index event.^106^ A second study of 517 patients did not identify any association between PM2.5, black carbon, or nitrogen dioxide, and ICH, and used a similar design comparing exposures 1-7 days prior to the hemorrhage to a referent set of days in the same month.^107^ PM2.5 and PM10 were not shown to associate with hemorrhagic stroke in a large Chinese study involving 69,399 subjects.^86^

#### Multiple Sclerosis

MS incidence as it relates to long-term air pollution exposure was assessed in one study of 6,203 patients, which did not show an association.^108^ Two studies, together comprising over 10,000 MS relapses, examined short-term PM10 exposure and the risk of MS-related hospitalization. There was a strong association between MS relapse and PM10 exposure within the first 1-3 days or one week prior to the index event.^109,110^ One case-crossover study examined short-term PM2.5 exposure and MS relapse, with an increased risk of relapse related to exposure 3 weeks prior.^111^ Two studies examined the short-duration effects of air pollutants (diesel exhaust or particulate matter versus filtered medical air) on serum markers of neuroinflammation within 24h of exposure (IL-6, TNF-alpha, BDNF, S100b, NSE, urinary VMA), and showed either inconsistent or no statistically significant differences between exposed and control groups.^86,112^ The pro-inflammatory association with PM10 exposure may relate to the chemokine receptor CCR6 in CD4+ T cells, found to be preferentially upregulated in MS patients after exposure.^113^

#### Headaches

Three studies examined recurrent headaches and pollutant exposures. The largest used a case-control design and leveraged electronic health records to examine 89,575 cases of migraine in California, with a higher frequency of migraine-specific urgent care visits for increasing average annual PM2.5 and NO2 levels.^114^ Two smaller studies, each with at least 18,000 patients, examined short-term pollutant exposures and visits for migraine, with statistically significant effects for PM2.5, PM10, NO2, O3, and CO exposures in adults, and PM2.5, CH4, SO2, NO2, and total hydrocarbon exposures in children.^115,116^

#### Parkinson’s Disease

Parkinson’s disease incidence as it relates to particulate matter exposure in the long-term has been examined by multiple studies. Three studies showed inconsistent or no significant association^117–119^, two showed association with PM10^120,121^ and one showed association with PM2.5.^122^ Among the most robust studies was a case-control study involving 38,475 PD patients from Ontario, where 2-year exposures for PM2.5, NO2, and O3, at a resolution of 1km x 1km were shown to associate with PD incidence (HRs ranged from 1.03-1.04).^122^ NO2 exposure was examined in five additional studies and its impact remains controversial, with increased concentrations noted in PD hotspots^121^, and two additional studies (combined 11,525 PD patients) suggesting association^123,124^, in opposition to two smaller studies (combined 1,496 PD patients) that did not show any statistically significant association.^117,120^

#### ALS

ALS incidence related to air pollutant exposure was studied with varying results. One study involving 917 ALS patients from a Dutch national registry showed an increased risk of ALS in patients with long-term exposure to PM2.5 and NO2, but another series of 52 patients examining PM10 exposure did not replicate these findings.^125,126^ Among patients with ALS and Parkinson’s disease, less controversial is the risk of disease aggravation in association with shorter-term exposure to PM2.5, which may lead to first-time admission and diagnosis of the underlying neurological condition.^127^

#### Neurodevelopment

Two studies were identified within our search strategy that examined neurodevelopment and pollutant exposure. Both studies examined over 500 children combined in urban settings in the United States and Mexico. Pollutant exposures in these studies were remarkably detailed, at resolutions of 1km x 1km to 10km x 10km, and outcomes examined included go-no go tasks or measures of neurocognitive performance. Both studies showed that prenatal PM2.5 exposure, particularly during the third trimester, after adjustment for various confounders, predicted poorer neurocognitive outcomes.^128,129^

## Discussion

Our scoping review has identified a wide body of literature pertaining to the impacts of climate change on neurologic disease. The results highlight many connections between the incidence and severity of neurologic disease and the effects of climate change and pollution. Perhaps the most striking finding of this review was how little is known regarding the effects of meteorological changes on neurologic illness, or on how the delivery of neurologic care should be adapted in the face of climatic events and changes.

This review has brought forward three key areas for expedited study owing to their potential for rapid change, large scale impact on neurologic health, and the potential for risk mitigation. Foremost, there is a priority to develop an understanding of emergent neurotropic infectious diseases. Diseases such as Zika virus, West Nile virus, tick-borne encephalitis, and COVID-19 have both the potential and precedence for very rapid spread across susceptible populations, with only a rudimentary understanding of their long-term effects on the nervous system. In the bulk of studies examined, we found evidence for vector incidence linked to climate, land use patterns, and human activity. An understanding of these factors and how they relate to disease vectors has given rise to novel strategies for risk mitigation, such vaccination programs, surveillance systems, and ecological defenses such as the Wolbachia bacteria in the case of Zika virus.^130,131^ In addition, as the effects of climate change intensify, migration and increased numbers of ‘climate refugees’ could result in larger populations to non-vector borne infections, such as COVID-19, and strategies to both study and mitigate these risks are needed.

Second, we identified an opportunity to enhance understanding of the risk of neurologic disease as it relates to airborne pollutant exposure. Specifically, multiple studies examined the degree of air pollution associated with cerebrovascular disease and dementia. Urban centers with high population density in developing nations are regions with the highest concentrations of airborne pollutants, and therefore represent a large population at risk for potentially preventable, chronic, neurologic diseases. An understanding of the pathophysiology of pollutants on the nervous system, as well as reduction and mitigation strategies, will be crucial to prevent increases in the incidence of these diseases.

Third, our work suggests that there is a potential unmet need in planning for the robust delivery of neurologic care in the face of ecological instability. We propose that tele-neurology may serve as a key solution to the disruptions that may occur due to climate changes in order to maintain continuity of neurologic care. The COVID-19 pandemic has highlighted how telemedicine can help patients obtain health-care access but could not otherwise access it.^132^ Further infrastructure, investment, and development of clinical skills and best practices in tele-neurology will be increasingly important to deliver care in a world with increasingly extreme weather events. Research into how to effectively manage patients with complex chronic neurologic conditions through tele-medicine is needed, as well as strategies for developing tele-neurology infrastructure in previously inaccessible regions.^133^

The present study has several limitations; first, we were limited to studies that were published in English or with suitable English translation. A risk of bias assessment for this scoping review was not performed, given the retrospective, observational nature of nearly all studies. Studies often relied on multi-variate modeling and were therefore biased by choice and measurement of the variables included in each of the models; differing modeling approaches often did not allow for direct comparison or meta-analysis of results. In addition, there are challenges unique to meteorological literature, including limitations on the spatial resolution of climate data, missing historical data, instrument-dependent effects, and limited understanding of true individual exposures due to behavioral variation. Modeling climactic variables’ effects on health outcomes often involves a process of selecting a lag time between exposure and outcome of interest, which can be arbitrary. There was significant variability in the results of the reviewed studies as differing lag times were often considered; a factor that limited inter-study comparison. Studies identified showed a geographic bias towards higher-income countries, whereas climate change is predicted to disproportionately affect those in developing nations. For example, our search strategy did not identify any studies from South America, and only five studies from Africa.

Practicing neurologists must consider the impact of climate change on each patient and their presentation. Akin to the social determinants of health, a changing climate cannot be overlooked as a key mediator of disease burden. The elderly patient with dysautonomia or MS and no access to air conditioning, the patient with symptoms of a small-vessel stroke in a city with increasing air pollution, or the patient with tick-borne encephalitis due to a vector never seen in their region previously, are all vignettes that will undoubtedly become more common. Their neurologic conditions are directly caused or exacerbated by the impacts of human-induced climate change, and we must acknowledge the burden of illness on those who bear little responsibility for greenhouse gas emissions. In doing so, our goal is to inspire action, research, and elicit true change to mitigate the risks of an already changing climate.

## Data Availability

All data produced in the present work are contained in the manuscript

## Supplementary Information

**Supplementary Table 1.** Studies of neuro-infectious diseases and their locations. Studies of neuro-infectious diseases associated with climate change, article type (primary or review), and the primary location of study, that were included in scoping review. TBE refers to tick-borne encephalitis. WNV refers to West Nile virus. JEV refers to Japanese encephalitis virus. DENV refers to Dengue virus.

| Study | Ref. | Organism | Location | Type |
| --- | --- | --- | --- | --- |
| Agier 2013 | 1 | TBE | Africa | Primary |
| Andersen 2017 | 2 | WNV, TBE | Global | Review |
| Andreassen 2012 | 3 | TBE | Europe | Primary |
| Bai 2013 | 4 | mosquito borne illness systematic review | Asia | Review |
| Baylis 2016 | 5 | JEV | Asia | Primary |
| Bi 2007 | 6 | JEV | Asia | Primary |
| Daep 2014 | 7 | flaviviruses (DENV, JEV, WNV) | Global | Review |
| Daniel 2018 | 8 | TBE | Europe | Primary |
| Daniel 2008 | 9 | TBE ixodes | Europe | Primary |
| Davis 2018 | 10 | WNV | North America | Primary |
| Dhimal 2015 | 11 | Dengue, lymphatic filariasis, JEV, visceral leishmaniasis, malaria | Asia | Review |
| Dobrotvorski 1999 | 12 | TBE | Asia | Primary |
| Gould 2009 | 13 | WNV, Chikungunya, rift valley fever, bluetongue virus | North America | Review |
| Harrigan 2014 | 14 | WNV | North America | Primary |
| Hsu 2008 | 15 | JEV | Asia | Primary |
| Jaenson 2018 | 16 | TBE | Europe | Review |
| Lindgren 2000 | 17 | TBE ixodes | Europe | Primary |
| Lindgren 2001 | 18 | TBE | Europe | Primary |
| Marini 2016 | 19 | Culex pipiens (mosquito) | Europe | Primary |
| Mazamay 2020 | 20 | Meningitis unspecified | Africa | Primary |
| Mihailovic 2020 | 21 | WNV, Anopheles hyrcapus | Europe | Primary |
| Molesworth 2003 | 22 | meningitis | Africa | Primary |
| Morin 2013 | 23 | WNV | North America | Primary |
| Murty 2010 | 24 | JEV | Asia | Primary |
| Paull 2017 | 25 | WNV | North America | Primary |
| Rainham 2005 | 26 | WNV | North America | Review |
| Randolph 2000 | 27 | TBE | Europe | Review |
| Russell 1998 | 28 | arbovirus | Australia | Review |
| Samy 2016 | 29 | TBE ixodes | Global | Primary |
| Shocket 2020 | 30 | WNV | North America | Primary |
| Singh 2020 | 31 | JEV | Asia | Primary |
| Skaff 2020 | 32 | Culex quinquefasciatus (mosquito) | North America | Primary |
| Soverow 2009 | 33 | WNV | North America | Primary |
| Sumilo 2007 | 34 | TBE | Europe | Primary |
| Sumilo 2008 | 35 | TBE | Europe | Primary |
| Tokarevich 2017 | 36 | Culex quinquefasciatus (mosquito) | Asia | Primary |
| Uejio 2012 | 37 | WNV | Africa | Primary |
| Vonesch 2016 | 38 | WNV, TBE | Europe | Review |
| Wang 2010 | 39 | WNV | North America | Primary |
| Zeman 2004 | 40 | TBE | Europe | Primary |
| Zhang 2016 | 41 | JEV | Asia | Primary |

